# Post-discharge health-related quality of life, cognitive function, disability, risk of post-traumatic stress disorder, and depression amongst the survivors of veno-venous extracorporeal membrane oxygenation (VV-ECMO) during the COVID-19 pandemic: a nested cohort study protocol

**DOI:** 10.1101/2024.08.22.24312450

**Authors:** Eunicia Ursu, Ana Mikolić, Sonny Thiara, Noah D. Silverberg, Denise Foster, William Panenka, Nishtha Parag, Mypinder S Sekhon, Donald E. G. Griesdale

## Abstract

**Background:** Veno-venous extra-corporeal membrane oxygenation (VV-ECMO) is a form of mechanical respiratory support for critically ill patients with severe acute respiratory distress syndrome (ARDS). Using a large intravenous line in a closed-circuit, blood is removed from the patient and passed through a hollow-fiber membrane where oxygen is added and carbon dioxide is removed. The oxygenated blood is then reinfused into the patient. Overt neurologic injury (ischemic stroke or intracerebral hemorrhage) occurs in approximately 20% of patients who receive VV-ECMO. However, it is unclear if there is additional unrecognized neurologic disability amongst patients who survive VV-ECMO. As such, we will perform a cohort study nested within our existing prospective study of patients who underwent VV-ECMO during the COVID-19 pandemic^1,2^. We expect to ascertain long-term patient reported and performance-based outcomes in greater than 60% of survivors of VV-ECMO. This study will provide important patient-centric long-term outcomes in contrast to the majority of existing studies of patients on VV-ECMO which focus solely on short-term survival.

**Methods and analysis:** We will include 39 patients who survived VV-ECMO and ascertain patient reported and performance-based outcomesthrough phone interviews. We will measure: i) Health-Related Quality of Life (HRQoL) using the EQ-5D-5L, ii) cognitive function using the T-MoCA Short, iii) disability using the World Health Organization Disability Assessment Scale (WHODAS) 2.0, iv) post-traumatic stress disorder (PTSD) using the Impact of Event Scale-6 (IES-6), and v) depression using the Patient Health Questionnaire – 9 (PHQ-9).

**Ethics and dissemination:** The results from the analysis of the study data will be disseminated through presentation of a scientific abstract at international conference, and submission of a manuscript in a peer-reviewed critical care medicine journal. The study ethical approval has been obtained from the University of British Columbia (UBC) Clinical Research Ethics Board (REB)(H21-00033) and the Vancouver Coastal Health Research Institute (V21-00033).

**Strengths and limitations of this study:**

➢ This study will provide important patient-centric long-term outcomes in different domains: symptoms, quality of life, functioning and cognition, in contrast to the majority of existing studies of patients on VV-ECMO during COVID-19 pandemic which focus solely on short-term survival.
➢ Describing the long-term outcomes in participants who had a documented neurologic injury while on VV-ECMO will bring new evidence-based data to potentially enhance the ELSO guidelines.
➢ Our study is a small, single centre design in the Western Region of Canada that may limit generalizability of results.
➢ Our sample size (majority are COVID-19 patients) will limit the ability to adjust for all relevant characteristics, and some relevant information is not available, in the exploratory analysis.

## Introduction

There have been over 760 million infections and over 6.8 million deaths globally since the start of the COVID-19 pandemic^3^. The acute hypoxemic respiratory failure due to COVID-19 led to a significant increase in the number of patients who needed mechanical respiratory support^4^.Veno-venous extra-corporeal membrane oxygenation (VV-ECMO) is a form of mechanical support for patients who develop severe ARDS refractory to mechanical ventilation therapies, including: pharmacologic paralysis, nitric oxide, and prone positioning^5^.

During VV-ECMO, venous blood is removed via a large catheter, pumped through a hollow-fibre membrane where oxygen is added and carbon dioxide is removed. The oxygenated blood is then reinfused into the patient^6^. Given the supporting data in patients with *severe* ARDS^7,8^, VV-ECMO has been used increasingly over the last decade, which accelerated with the COVID-19 pandemic^5^. Guidelines for the use of ECMO in patients with COVID-19 have been published by the Extracorporeal Life Support Organization (ELSO)^9^. However, the use of VV-ECMO has potentially expanded beyond the indications previously studied to include patients with *less severe* ARDS^10^. This is important as patients appear to be at high risk of neurologic injury while receiving VV-ECMO, including: cerebral infarct, intracerebral hemorrhage, and seizures^11–13^. Historical cohort studies reported among adult patients with acute respiratory failure undergoing VV-ECMO an incidence of neurologic injury ranges from 7 to 50%^14^. Thus, the purported benefit of VV-ECMO may not outweigh the risks of neurologic injury, particularly as the use of VV-ECMO is being expanded to include patients with *less severe* ARDS^15^.

Beyond the risk of acute neurologic injury, VV-ECMO may have relatively subtle but clinically important adverse effects on long-term functioning, but data addressing long-term effects are sparse and conflicting.There are several historical cohort studies with conflicting results in terms of Health-Related Quality of Life (HRQoL) and cognitive function^16–18^. As such, there is a paucity of data examining long-term outcomes in patients undergoing VV-ECMO for ARDS during the COVID-19 pandemic.

In our previous prospective cohort study of 59 patients (with 50 patients, representing 85% of total patients, that had respiratory failure secondary to COVID-19), who underwent VV-ECMO during the COVID-19 pandemic^1,2^, 12 patients (20%) suffered an acute neurologic injury (ischemic stroke or intracerebral hemorrhage) while receiving VV-ECMO in the ICU. This proposed nested cohort study aims to assess long-term neurologic outcomes in hospital survivors from the prospective cohort study.

Specifically, we will assess survivors in the following domains: i) Health-Related Quality of Life (HRQoL), ii) cognitive function, iii) disability, iv) post-traumatic stress disorder (PTSD), and v) depression.

## Methods

### Study design and population

This will be a nested cohort study of patients who received, and were liberated, from VV-ECMO during the COVID-19 pandemic at Vancouver General Hospital (VGH) between April 2020 and November 2021^1,2^. The original cohort study of 59 patients examined biomarkers of neurologic injury amongst patients who received VV-ECMO (the UBC REB #H21-00033). This current study will examine the 39 of 59 (66%) of patients who were liberated from VV-ECMO and survived to ICU discharge. **Figure 1** shows the study flow diagram.

**Figure 1.**
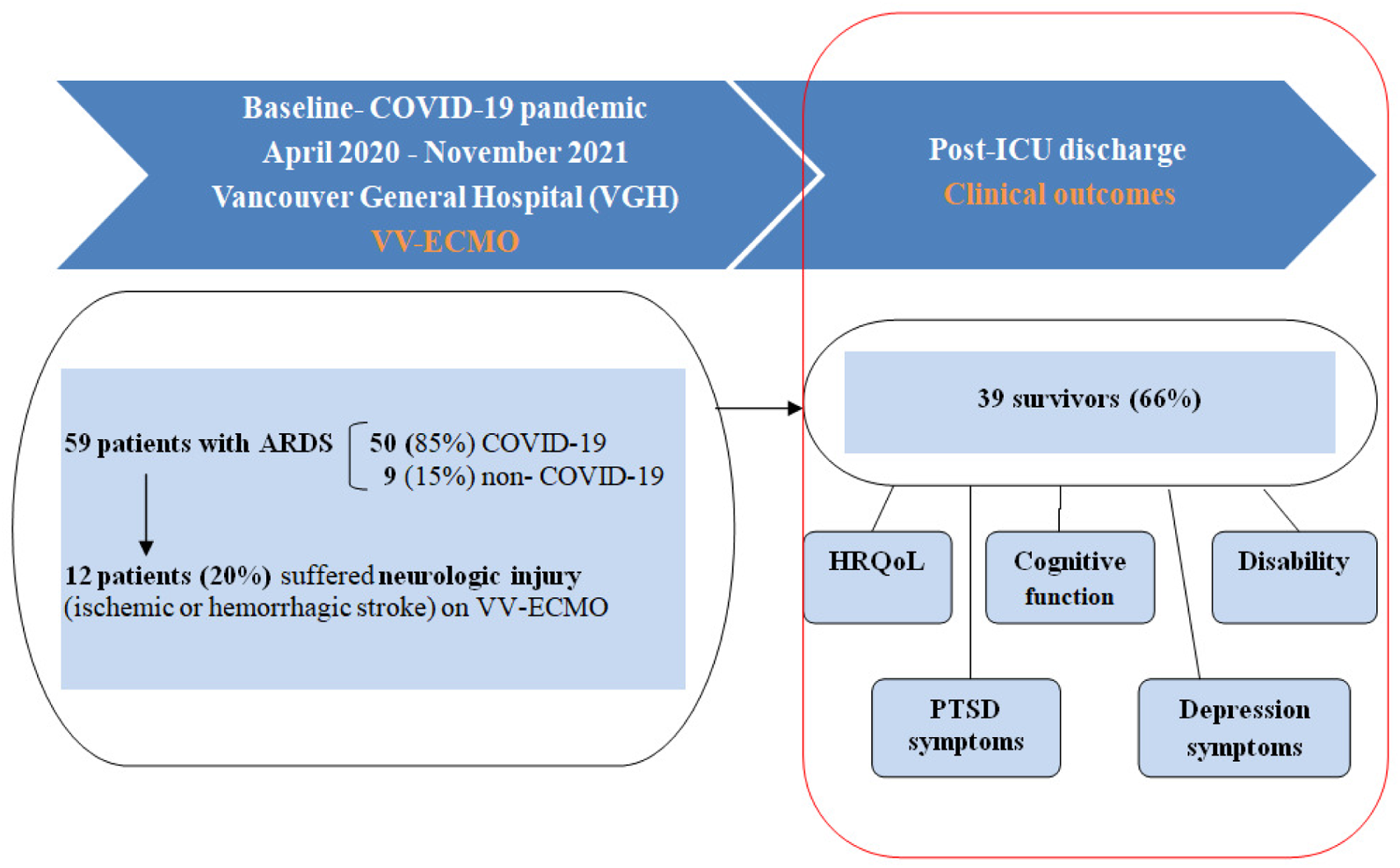
Study flow diagram

### Baseline data collection and measures

Demographic and health information were collected as part of the parent study using a Research Electronic Data Capture (REDCap)(H14-00930) database^19^.Acute clinical data including VV-ECMO related central nervous system injury (ischemic stroke or intracerebral hemorrhage) along with multi-modal biomarkers of neurologic injury (neurofilament light, glial fibrillary acidic protein, and phosphorylated-tau 181) prior to, and 1-hour, 24-hours and 7-days following initiation of VV-ECMO were also collected during the ICU stay.

### Long-term neurologic outcome

We will collect data on clinical outcomes using a combination of patient reported and performance-based outcomes. Due to the ongoing COVID-19 pandemic, particularly when working with potentially vulnerable patients, we will contact the survivors and administer these tools by either phone or video-conferencing (e.g., Zoom) interview. We will use an abbreviated 30 -minute battery. This project will be implemented according to the time schedule shown in Table 1. Based on the proposed assessment dates, we anticipate a follow-up range of between 15 and 30-months.

**Table 1.**
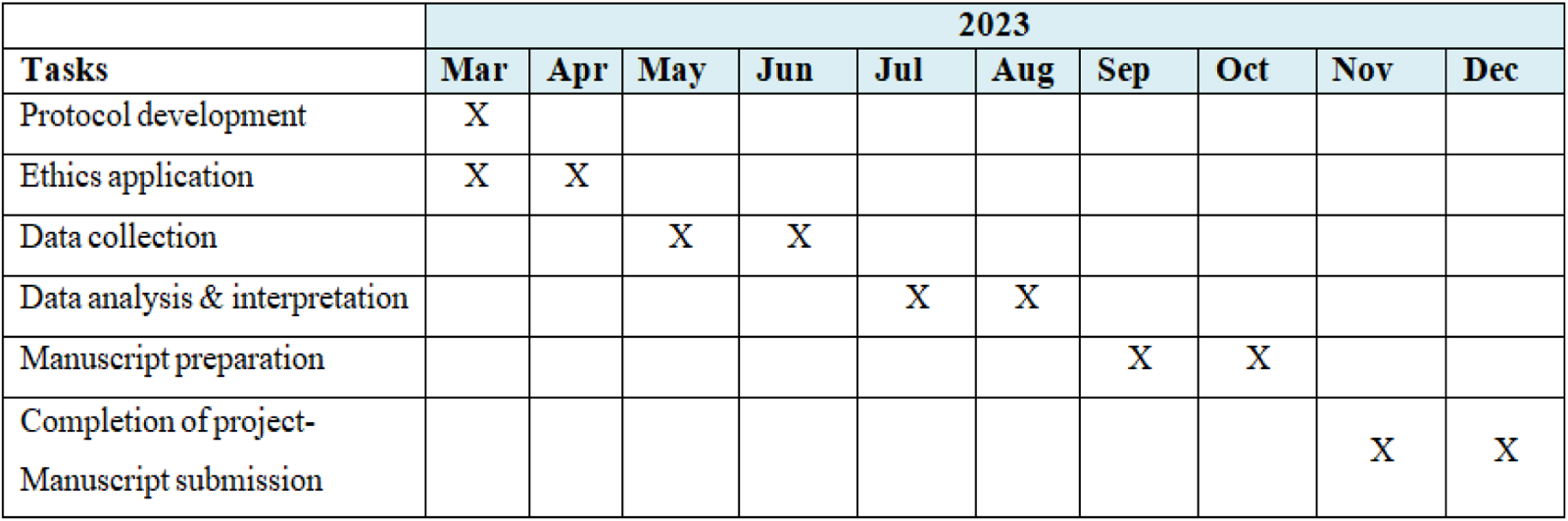
Schedule of the Proposed Study.

| Tasks | 2023 |  |  |  |  |  |  |  |  |  |
| --- | --- | --- | --- | --- | --- | --- | --- | --- | --- | --- |
|  | Mar | Apr | May | Jun | Jul | Aug | Sep | Oct | Nov | Dec |
| Protocol development | X |  |  |  |  |  |  |  |  |  |
| Ethics application | X | X |  |  |  |  |  |  |  |  |
| Data collection |  |  | X | X |  |  |  |  |  |  |
| Data analysis & interpretation |  |  |  |  | X | X |  |  |  |  |
| Manuscript preparation |  |  |  |  |  |  | X | X |  |  |
| Completion of project-Manuscript submission |  |  |  |  |  |  |  |  | X | X |

### Clinical outcomes

The research team will assess long-term outcomes that focus on the following domains: i) HRQoL, ii) cognitive function, iii) disability, iv) PTSD, and v) depression in patients undergoing VV-ECMO for ARDS during COVID-19 pandemic.

### Study Instruments

The following validated instruments will be administered: EuroQoL-5D-5L (EQ- 5D-5L)^20^, the telephone Montreal Cognitive Assessment (T-MoCA Short)^21^, the World Health Organization Disability Assessment Schedule (WHODAS 2.0)^22^, the Impact of Event Scale-6 (IES-6)^23^, and the Patient Health Questionnaire – 9 (PHQ-9)^24^. The outcome instruments and measures are based on international consensus recommendations^25,26^, and were selected in consultation with the UBC Clinical Outcomes for Brain Research - Assessment Hub (COBRAH) (https://cobrah.psych.ubc.ca). **Table 2** summarizes our study instruments and measures.

**Table 2.** Instruments and measures of Proposed Study.

| Instrument | Domain | Description | Generic<sup>1</sup><br>or brain<br>injury<br>specific | Domains | Versions<br>- Length<br>(items) | Duration<br>(approx.<br>in<br>minutes) |
| --- | --- | --- | --- | --- | --- | --- |
| EQ-5D-5L<sup>2</sup> | **Health Related**<br>**Quality of Life** | Self-rated<br>health<br>status | Generic | Mobility<br>Self-care<br>Usual activities<br>Pain/ discomfort<br>Anxiety/depression | 5 (more<br>response<br>levels)<br>+VAS | <5 min. |
| T-MoCA<br>Short | **Cognitive**<br>**function** | Brief<br>telephone<br>screening<br>for cognitive<br>impairment | / | Attention, verbal<br>learning and<br>memory, executive<br>functions/language,<br>and orientation:<br>6 orientation items,<br>60-second "F"<br>fluency,<br>5-word immediate<br>and delayed<br>memory | 12 points | 5 min. |
| WHODAS<br>2.0 | **Functional**<br>**outcome** | Disability<br>assessment | Generic | Cognition<br>Mobility<br>Self-care<br>Getting along<br>Life activities<br>Participation | 12 | 5 min. |
| IES-6 | **Posttraumatic**<br>**stress** | Screening<br>for PTSD:<br>subjective<br>stress<br>caused by<br>traumatic<br>events | / | Intrusion<br>Avoidance<br>Hyperarousal | 6 | <5min |
| PHQ-9 | **Depressive**<br>**symptoms** | Screening<br>for<br>depression/<br>severity of<br>depression<br>symptoms | / | Symptoms | 9 | <5min |
| EQ-5D-5L= EuroQoL-5D-5L; VAS = visual analogue scale; WHODAS 2.0= World Health Organization Disability Assessment Schedule 2.0; T-MoCA= Telephone Montreal Cognitive Assessment; IES-6= Impact of Event Scale-6; PTSD= Post-Traumatic Stress Disorder; PHQ-9=Patient Health Questionnaire– 9 |  |  |  |  |  |  |

<sup>1</sup>for different health conditions; <sup>2</sup>included in the core outcome set.

#### EQ-5D-5L

The EQ-5D-5L is a six-item questionnaire measuring health-related quality of life^20^. It assesses five domains: mobility, self-care, usual activities, pain/discomfort and anxiety/depression, each containing five responses from ‘no problem’ to ‘extreme’ problems or ‘unable to’. There is also a question of overall quality of life using a visual analogue scale from 0 (poor state of health) to 100 (good state of health)^20^. The EQ-5D- 5L will be used to evaluate quality of life in VV-ECMO survivors at the time of follow-up.

#### T-MoCAShort

The T-MoCA Short is a brief (5-minute) instrument for assessing cognition over the telephone ^21,27^. It is a validated screening tool for cognitive function in stroke patients^27^. T-MoCA Short assesses attention, verbal learning and memory, executive functions/language, and orientation. A total score (ranges from 0 to 15) of below the cutoff score of 12 will be interpreted as potential mild cognitive impairment (MCI)^28^.

#### WHODAS 2.0^22^

The WHODAS 2.0 (12-item interview version) will be used to assess disability. This instrument covers 6 domains of functioning: cognition, mobility, self-care, getting along, life activities and participation. It has been used across a variety of health conditions, including COVID-19^29^.Each item is rated from 1-’no difficulty’ to 5-’extreme difficulty or cannot do’ based on how much difficulty a patient has had with an activity over the past 30 days because of a health condition. Disability severity is based on the total sum score, with higher scores indicating greater global disability^22^. When participants will not be able to complete the WHODAS themselves, their proxy (e.g., relative, carer, etc.) will complete similar questions administered as a self-report measure (survey) titled the WHODAS 2.0 12-item proxy.

#### IES-6

The IES-6 is a reliable and valid research instrument for PTSD amongst survivors of ARDS^23^.The IES-6 is a 6-item short version of the Impact of Event Scale-Revised (IES-R)that measures symptoms of post-traumatic stress, including intrusive thoughts, hyperarousal, and avoidance of reminders of a traumatic event.Itis a recommended measurement tool for ICU survivors in general^23,30^, and achieved a sensitivity of 88% and specificity of 85% for detecting PTSD in ARDS survivors^23^. IES-6 score will be calculated as the mean of the six items which have a response scale of: 0 =’not at all’, 1 = *‘*little bit’, 2 = ‘moderately’, 3 = ‘quite a bit’, and 4 = ‘extremely’. The cut-off of 1.75 (mean of the 6 items) will indicate the presence of clinically significant PTSD symptoms^23^.

#### PHQ-9 instrument

The PHQ-9 is a self-reported measure of depression symptom severity over the prior two weeks^24^.PHQ-9 score ≥ 10 has 88% sensitivity and 88% specificity for major depressive disorder. The nine items assessed are: anhedonia, depressed mood, insomnia or hypersomnia, fatigue or loss of energy, appetite disturbances, guilt or sense of worthlessness, diminished ability to think or concentrate, psychomotor agitation or retardation, and suicidal thoughts. In each response category, scores range from 0 to 3 representing ‘not at all’, ‘several days’, ‘more than half the days’, and ‘nearly every day’, respectively. The total score (sum of the 9 items)>=10suggests presence of depressive symptoms^24^.

### Withdrawal from study

Either the participant or the Temporary Substitute Decision Maker (TSDM) can request withdrawal from the study. Once withdrawn from the study, no further study procedures or evaluations will be performed. We will attempt to obtain permission to document the reason for withdrawal and to collect participant outcomes. Any data captured prior to the withdrawal of consent will be retained if consent is obtained to do so.

### Patient participation at post-discharge clinical outcomes

The phone and online interviews will be administered by the COBRAH clinical research coordinators and research assistants with training on standardized patient-reported questionnaires administration of neurological/neuropsychological tests, under the supervision of a neuropsychologist. The COBRAH personnel have clinical and research experience with the proposed outcome measures^31–34^.

### Sample size and statistical analysis plan

#### Sample size

We aim to examine the 39 of 59 (66%) of patients who were liberated from VV-ECMO and survived to ICU discharge between April 2020 and November 2021.We deem this study feasible if we are able to obtain these measures in >60% of patients.

#### Statistical analysis plan

We will present descriptive statistics on the baseline retrospective data collection (e.g., age, sex, Acute Physiology Chronic Health Evaluation score -APACHE II, days spent on ECMO support, length of stay on ICU) for each of the scores across all patients. We will report medians with interquartile ranges for continuous distributed variables. Categorical variables will be presented with percentages. For each of the EQ- 5D-5L, WHODAS 2.0, T-MoCA Short, IES-6 and PHQ-9scores we will describe outcomes in terms of distributions, medians with interquartile ranges, and percentages of participants meeting threshold criteria for impairment. Finally, we will plot the glial fibrillary acidic protein levels over time in those patients who have either PTSD or depression.

A two-tailed P-value < 0.05 will be considered statistically significant. We will use R version 4.2.2^35^ for all statistical analyses.

## Ethical Considerations and Informed Consent

### Ethical Considerations and informed consent

Ethical approval has been obtainedfrom the UBC REB(H21-00033) and the Vancouver Coastal Health Research Institute (V21-00033).Consent will be obtained from either the participant or their substituted decision maker. Substitute decision maker consent will be obtained in cases where the participant is unable to participate in the outcome assessment interviews. Consent for outcome assessments will either be obtained while participants are in hospital or at time of the post-discharge follow-up for the EQ-5D-5L as part of the original cohort study. Upon contact, we will re-consent participants to assess the T-MoCA Short, WHODAS 2.0, IES-6, PHQ-9. A gift card will be provided to compensate the participant/TSDM for their time. Information related to this is provided in the consent form.

### Assessment of Depression

As the PHQ-9 includes a question about suicidal thoughts, a standardized Distress and Suicidal Protocol^36^ will be used to clarify and triage suicide risk. This protocol will be followed in cases where an individual reports experiencing suicidal thoughts “More than half the days” or “Nearly every day” on the PHQ-9, and involves administering the Columbia-Suicide Severity Rating Scale-Screener (C-SSRS) to determine an action matched to the level of suicide risk, which ranges from providing coping resources to referring for medical care or dialing 911(**Figure 2**).

**Figure 2.**
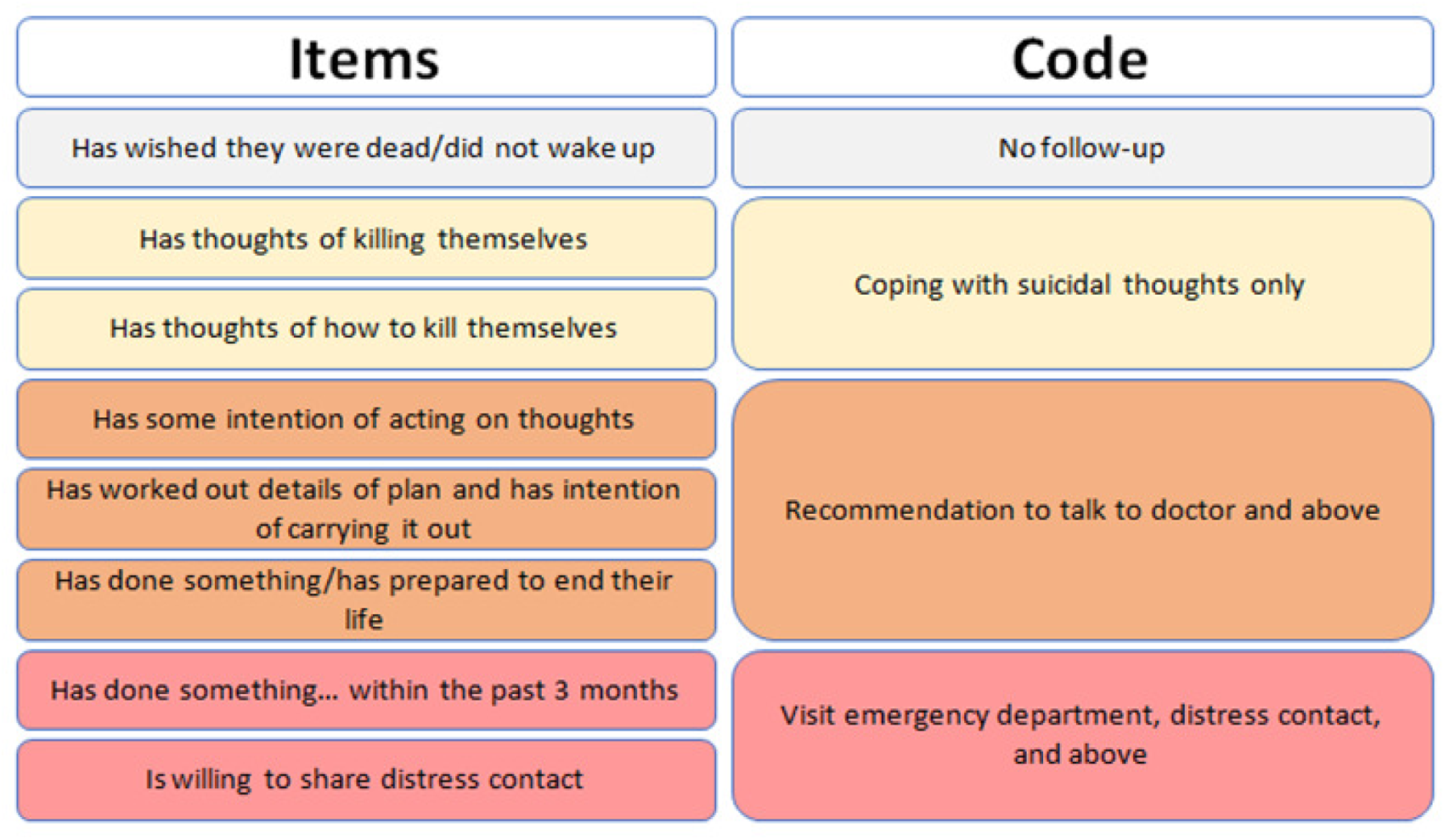
Standardized Distress and Suicidal Protocol

### Potential threats for ascertainment of patient reported performance-based outcomes

VGH is a quaternary care center and many of our patients are admitted by transfer from rural British Columbia. Therefore, to decrease patient attrition and improve feasibility, we chose outcomes measures which can be completed remotely, by either telephone or video link (e.g. Zoom). Our team has extensive experience with our chosen outcome battery^31–34^as well as experience with recruitment and retention of patients in remote outcome studies^37^. Finally, we have developed a reimbursement and remuneration strategy to help support participants so that outcomes can be assessed.

## Data Availability

All data produced in the present study are available upon reasonable request to the authors.

## Abbreviations and Definitions of Terms

ARDS: Acute Respiratory Distress Syndrome
COVID-19: Coronavirus Disease 2019
ELSO: The Extracorporeal Life Support Organization
EQ-5D-5L: EuroQoL-5D-5L
HRQoL: Health-Related Quality-of-Life
IES-6: Impact of Event Scale-6
PTID: Patient Identifying Number
PTSD: Post-Traumatic Stress Disorder
PHQ-9: The Patient Health Questionnaire – 9
REDCap: Research Electronic Data Capture
T-MoCA: The telephone Montreal Cognitive Assessment
TSDM: Temporary Substitute Decision Maker
VV-ECMO: Veno-Venous Extracorporeal Membrane Oxygenation
WHODAS 2.0: World Health Organization Disability Assessment Scale 2.0

